# Multi-tissue transcriptome-wide association study identifies 29 risk genes associated with attention-deficit/hyperactivity disorder

**DOI:** 10.64898/2026.02.16.26346287

**Authors:** Sarina Abrishamcar, Qile Dai, Jingjing Yang, Anke Hüls, Michael P. Epstein

## Abstract

**Background:** Attention-deficit/hyperactivity disorder (ADHD) is a common heritable neurodevelopmental disorder, affecting ∼7 million children (11.4%) in the U.S. However, ADHD’s underlying genetic architecture remains largely unknown. Transcriptome-wide association studies (TWAS), which integrate expression quantitative trait loci (eQTL) and GWAS summary data, can identify differentially expressed risk genes underlying complex phenotypes. Here we conduct a TWAS of ADHD using expression data from multiple brain tissues to improve understanding of the complex genetic architecture underlying this psychopathology.

**Methods:** We applied the TWAS framework OTTERS to train multiple gene expression imputation models using cis-eQTL summary statistics from MetaBrain for three brain regions: cortex (n=2,683), basal ganglia (n=208), and cerebellum (n=492), and GWAS summary statistics from the most recent meta-analysis of ADHD (n=225,534; case fraction =0.17). We further conducted fine-mapping, colocalization analysis, and functional enrichment analysis.

**Results:** We identified 29 significant TWAS risk genes for ADHD (11 in cortex, 4 in basal ganglia, and 14 in cerebellum). Six genes appear novel for ADHD (*MPL, C1orf210, MDFIC, NKX2-2, FAM183A, HIGD1A)* while four genes were previously implicated in autism spectrum disorder (*XRN2*, *KIZ*, *NKX2-4*, *NKX2-2*). Pathway analysis indicated cortex and basal ganglia were enriched for neurodevelopmental pathways and regulation of cell development, and the protein-protein interaction network was statistically significant (p=1.12E-04).

**Conclusion:** This multi-tissue TWAS refines the genetic architecture of ADHD by identifying genes whose genetically regulated expression is associated with risk, including six candidates not previously linked to ADHD. Together, these findings provide novel insights for potential targets in translational research and drug discovery.

## Introduction

Attention-deficit/hyperactivity disorder (ADHD) is an increasingly common neurodevelopmental disorder, affecting approximately 7 million children (11.4%) in the U.S. (1). Between 2016 and 2022, the number of children who had ever received an ADHD diagnosis increased by 1 million (1). Additionally, ADHD often co-occurs with other childhood psychopathologies such as learning disorders, anxiety or depression, and behavioral or conduct problems (1). Together these factors have been associated with missing key developmental milestones, decreased school performance, and decreased productivity in adulthood, highlighting the public health significance of ADHD and the importance of elucidating its underlying biological mechanisms. ADHD is one of the most heritable psychiatric conditions, with a recent meta-analysis of twin studies estimating the heritability to be between 77% and 88% (2). Two large-scale genome-wide association studies (GWAS) meta-analyses conducted in 2018 (N=20,183 cases, N=35,191 controls) and 2023 (N = 38,691 cases, 186,843 controls) identified 12 and 27 risk loci associated with ADHD, respectively (3,4). SNP-based heritability estimates based on such GWAS data range from 0.14 to 0.22 (3,4).Together, these findings suggest that ADHD is highly polygenic and implicates thousands of common variants in its etiology.

Despite these recent GWAS findings, the underlying genetic architecture of ADHD remains largely unknown. Many of the associated variants from these GWAS are located in non-coding regions, making functional interpretation challenging (5). Non-coding SNPs are often implicated in complex disease and are hypothesized to influence gene expression (6). Transcriptome-wide association studies (TWASs) integrate expression quantitative trait loci (eQTL) from gene expression data with GWAS summary data to identify genes that increase risk of complex diseases like ADHD through genetic regulation of the transcriptome (7–9). While previous TWAS of ADHD have identified putative risk genes, these studies have primarily focused on cortex tissue and failed to consider additional brain tissues (like cerebellum and basal ganglia) (10,11). Moreover, these existing TWAS used expression data from individual-level datasets like GTEx that has relatively small sample sizes rather than larger summary-level eQTL datasets available from resources like MetaBrain with larger sample sizes. TWAS methods now exist that can accommodate summary-level eQTL reference data for predicting genetically regulated gene expressions (GReX) that will be associated with the phenotype of interest, with results indicating that the increased sample size of such larger reference datasets increases TWAS power relative to smaller individual-level reference datasets (12,13).

Here we apply the Omnibus Transcriptome Test using Expression Reference Summary Data (OTTERS) framework to conduct a multi-tissue TWAS of ADHD (13). OTTERS leverages five polygenic risk score (PRS) methods to train gene expression imputation models using summary-level eQTL data and then performs downstream association testing by integrating these trained gene expression imputation models with the summary-level GWAS data.

Specifically, we integrate ADHD GWAS summary statistics from the Psychiatric Genomics Consortium (n=225,534; case fraction =0.17) with summary eQTL data from MetaBrain for three functionally relevant brain regions (cortex, cerebellum, and basal ganglia) that have been consistently implicated in the etiology of ADHD (4,14). Our objectives are to 1) identify genes whose genetically regulated expression is associated with ADHD risk, 2) evaluate the robustness of associations across brain regions, and 3) provide further insight into the genetic architecture underlying ADHD.

## Methods

### ADHD GWAS summary data

We used ADHD GWAS summary statistics from the 2023 Psychiatric Genetics Consortium meta-analysis, which included 38,691 ADHD cases and 186,843 controls, for a total sample size of 225,534 (case fraction=0.17) (4). These summary statistics were derived from GWAS meta-analyses using data from iPSYCH (25,895 cases; 137,991 controls) (15,16), deCODE genetics (8,281 cases; 137,993 controls), and data published from ten ADHD cohorts from the Psychiatric Genetics Consortium (4,515 cases; 186,843 controls) (3). Analysis was restricted to participants from European ancestry. GWAS was performed separately for iPSYCH1 and iPSYCH2 using dosage data and additive logistic regression, adjusting for the first ten principal components. GWAS for deCODE samples was conducting using dosage data and logistic regression, adjusting for sex, year or birth, and country of origin. To adjust for inflation due to population stratification and cryptic relatedness, test statistics were divided by the inflation factor estimated from LD score regression. GWAS summary statistics from the previous PGC ADHD GWAS were used for the ten ADHD cohorts(3). Meta-analysis of variants with imputation quality > 0.8 and MAF > 0.01 was conducted using fixed effects standard error meta-analysis with METAL (17). Variants that had an effective sample size greater than 60% were retained in the final summary statistics (6,774,244 variants). The study was approved by the local scientific ethics committees and institutional review boards, and all participants provided informed consent, as described in Demontis et.al. (4).

### MetaBrain eQTL data

We used genome-wide cis-eQTL summary statistics from MetaBrain for three brain regions, restricted to European ancestry: cortex (2,683 samples), basal ganglia (208 samples), and cerebellum (492 samples) (14). These summary data were generated using 8,613 bulk brain RNA-seq samples and 6,518 genotype samples across 14 publicly available datasets from seven major brain regions. Cis-eQTLs were defined as statistically significant (q-value <0.05) combinations of SNPs and genes within a 1 Mb window of the transcription start site. We used the GTEx V8 whole genome sequencing (WGS) data for European ancestry to construct the LD reference panel (18). The study was approved by the local scientific ethics committees and institutional review boards, as described in de Klein et.al. (14).

### Transcriptome-wide association study

We employed the OTTERS (Omnibus Transcriptome Test using Expression Reference Summary Data) framework to conduct two-stage TWAS using summary-level reference eQTL and GWAS data. The OTTERS framework has been described in detail in Dai et.al. (13,19). Briefly, in Stage 1, OTTERS leverages five PRS methods to train genetically regulated expression (GReX) regression prediction models where cis-eQTL coefficients were estimated for predicting the target gene expressions, using cis-eQTL summary-level data and an external LD reference panel of the same ancestry. These cis-eQTL coefficients are used as variant weights, referred to as cis-eQTL weights, to conduct gene-based association tests. These PRS methods include pruning and thresholding with p-value thresholds of (i) 0.05 (P0.05) and (ii) 0.001 (P0.001) (20), (iii) frequentist LASSO regression model (lassosum) (21,22), (iv) nonparametric Bayesian Dirichlet process regression model (SDPR) (23,24), and (v) Bayesian multiple linear regression model with continuous shrinkage prior (PRS-CS) (25).

In Stage 2, OTTERS performs gene-based association testing for each training method using the cis-eQTL weights estimated from Stage 1 and summary-level GWAS data. This results in a set of five TWAS p-values for each training method. OTTERS then conducts an omnibus test using the aggregated Cauchy association omnibus test (ACAT-O), which aggregates p-values using the Cauchy distribution into a final OTTERS p-value (26). The ACAT-O test is widely used in hypothesis testing when combining multiple statistical methods for the same hypothesis and has been shown to improve power while controlling for the type I error rate (26). We adjusted the OTTERS TWAS p-values using the genomic control factor to account for potential inflation. Finally, we corrected for multiple testing using the Bonferroni threshold for the number of genes tested per tissue (a =2.7E-06).

### Fine-mapping TWAS risk genes

To identify independent risk genes, we conducted fine-mapping on genomic regions containing more than one significant TWAS risk genes using GIFT (gene-based integrative fine-mapping through conditional TWAS) (27). First, GIFT performs conditional joint TWAS analysis that tests each gene-trait association while conditioning on GReX of neighboring genes. Next, GIFT conducts fine-mapping by conducting a likelihood ratio test by comparing the full joint model to the reduced model excluding each candidate gene to generate conditional p-values. This yields frequentist p-values for fine-mapped gene prioritization. We performed GIFT analysis using the cis-eQTL weights estimated by each of these considered five PRS models. As a set of five GIFT p-values were obtained for each gene in the test region, we conducted ACAT-O to derive a final GIFT p-value for each genomic region. The GIFT p-value was corrected for multiple testing using False Discovery Rate (FDR) correction with respect to the number of genomic regions.

### Colocalization analysis

We conducted Bayesian colocalization using the SuSIE-based method implemented in the coloc R package (28) on all significant TWAS genes. For each gene-tissue pair, COLOC was first used to derive credible sets of putative causal variants for the eQTL signal, which were then compared against COLOC-derived GWAS credible sets in a pairwise framework. COLOC-based colocalization requires genes that have a detectable eQTL signal and sufficient SNP overlap between the eQTL and GWAS summary statistics. COLOC did not identify credible sets if there was insufficient SNP overlap or weak eQTL signal and therefore colocalization results are not reported for all genes. Posterior probabilities (PP) were computed for five hypotheses (H0-no-association, H1-GWAS-only, H2-eQTL-only, H3-independent-signals, or H4-shared-causal-variant), with evidence for strong colocalization defined as PP.H4 > 0.8 and moderate colocalization as PP.H4 >0.5.

### Functional enrichment analysis

We conducted pathway enrichment analysis on all significant TWAS risk genes to identify potential biological pathways that may be overrepresented. We used the enrichGO() function from the clusterProfiler R package to conduct Gene Ontology (GO) enrichment analysis (29). We removed redundant terms using the simplify() function at a similarity threshold of 0.8. We also utilized the STRING database to assess potential protein-protein interaction (PPI) networks among the identified risk genes (30).

## Results

### eQTL architecture of OTTERS prediction models

To characterize the underlying eQTL architecture of the OTTERS prediction models, we summarized the number of genes with at least one non-zero weight and the median number of estimated eQTLs per gene for each method and tissue (Table S1). Across tissues, all methods produced a similar number of testable genes (range: 11,503-18,362). SDPR, PRScs, and lassosum methods had higher median numbers of estimated eQTLs per gene (2,740-3,379 eQTLs per gene) ,whereas the more stringent P+T methods produced substantially sparser prediction models (5-406 eQTLs per gene).

### Transcriptome-Wide Association Study

Using OTTERS, we performed TWAS analyses of ADHD based on summary-level cis-eQTL reference data from three relevant brain tissue (cortex, cerebellum, and basal ganglia) and GWAS summary data of ADHD. We observed moderately inflated lambda values across all three tissues (Figures S1-S6) in line with the original ADHD GWAS meta-analysis (inflation factor of 1.30) which the authors demonstrated to be mostly explained (90%) by polygenicity (4). Aiming for conservative inference, we adjusted the omnibus OTTERS p-value by the genomic control factor. Post adjustment, we identified 29 significant TWAS risk genes of ADHD with p-values less than the Bonferroni threshold (a = 2.7E-06), including 11 in cortex, 22 in cerebellum, and 4 in basal ganglia (Figure 1; Table 1). Figure 2 shows overlap of identified risk genes across brain tissues considered. We observed that *MPL* was an ADHD risk gene identified across all three brain tissues. Meanwhile, we found that *TIE1*, *IPO13*, *PTPRF*, and *MED8* were risk genes identified in both cortex and cerebellum while *NKX2-2* was a risk gene identified in both the basal ganglia and cerebellum.

**Figure 1:**
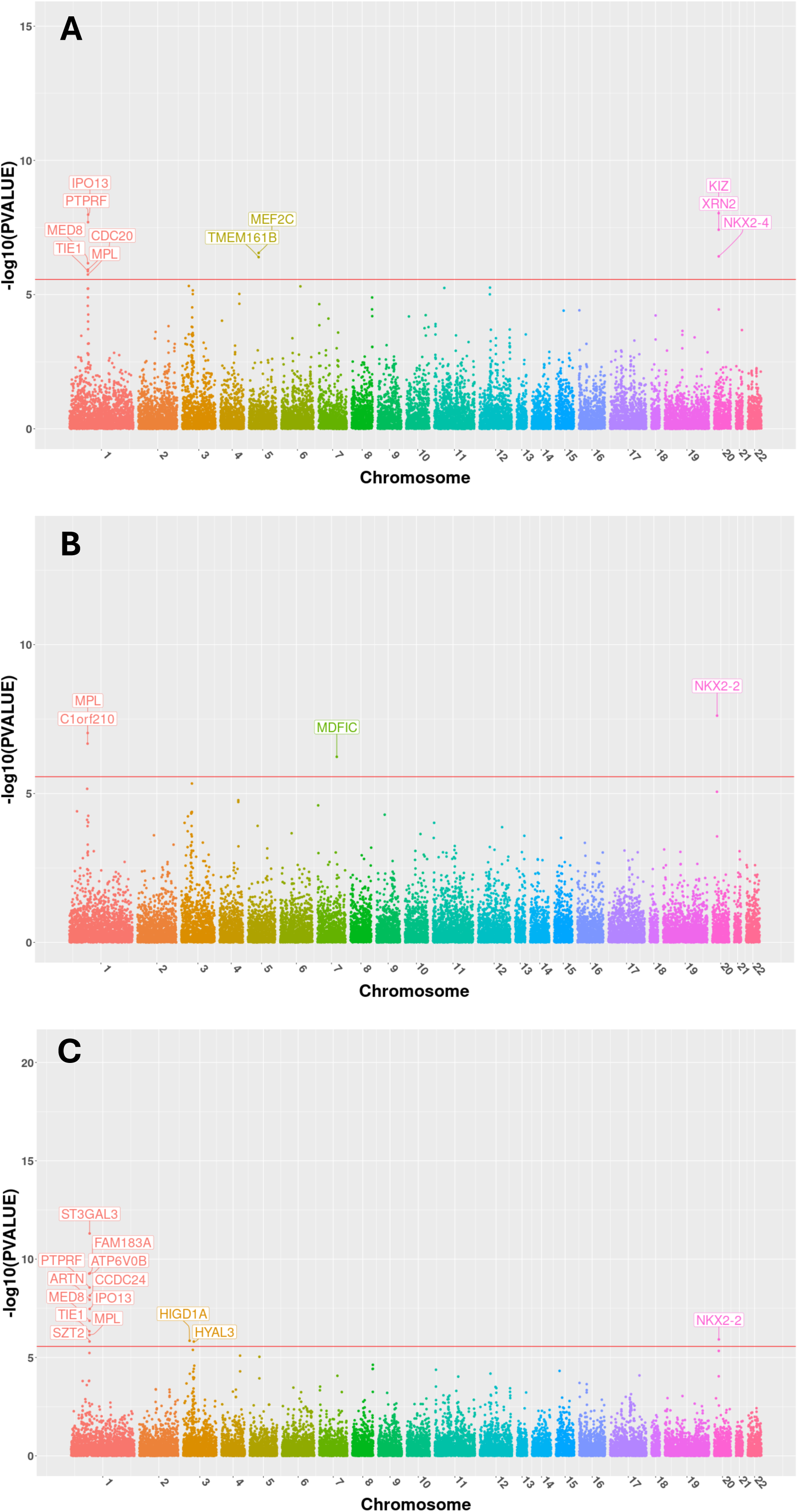
Manhattan plots of ADHD TWAS results across brain tissues. Manhattan plots show TWAS results for ADHD in A) cortex, B) basal ganglia, and C) cerebellum. Labeled genes surpass the genome-wide significance threshold, defined by a Bonferroni threshold of 2.7E-06.

**Figure 2:**
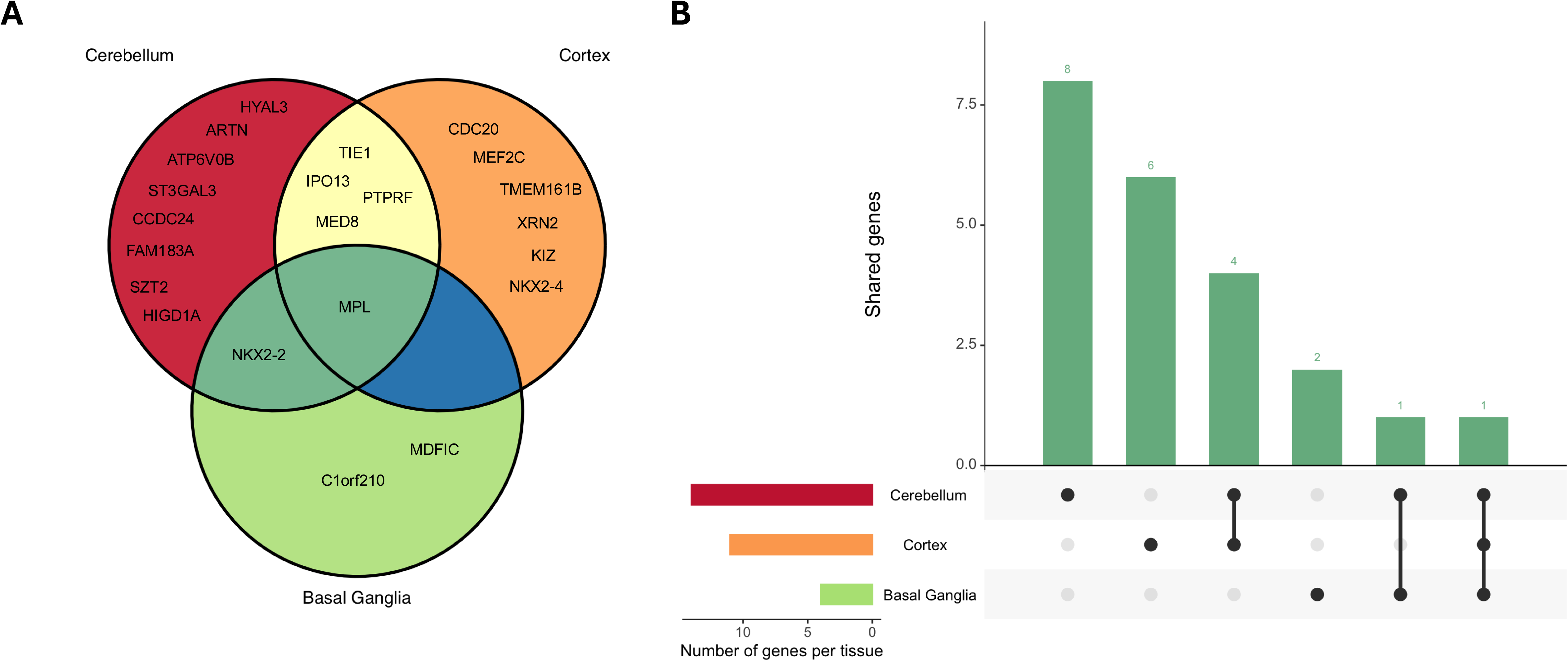
Overlap of significant ADHD TWAS genes across brain tissues. A) Venn diagram showing overlap of TWAS-significant genes across cortex, basal ganglia, and cerebellum. B) UpSet plot summarizing the number of shared and tissue-specific TWAS-significant genes across tissues.

**Table 1:**
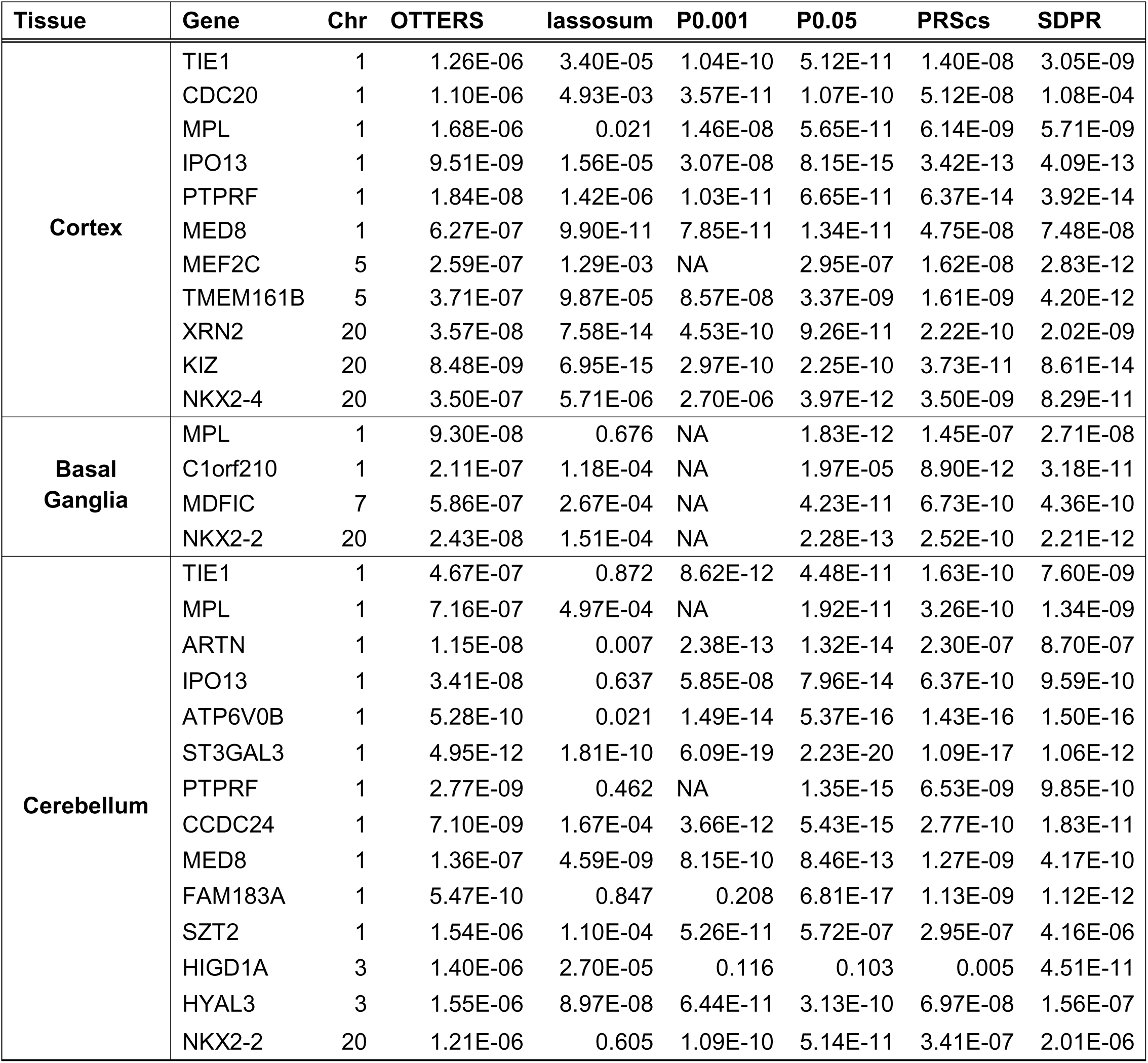
Significant OTTERS TWAS Results for ADHD. Shown are genes whose genetically related expression was significantly associated with ADHD risk across cortex, basal ganglia, and cerebellum at a Bonferroni threshold of 2.7E-06. For each tissue-gene pair, association *P* values are reported for the omnibus OTTERS test and for individual prediction methods. Missing values, noted by NA, indicate that a given method did not yield a valid prediction model for that gene-tissue pair.

| Tissue | Gene | Chr | OTTERS | lassosum | P0.001 | P0.05 | PRScs | SDPR |
| --- | --- | --- | --- | --- | --- | --- | --- | --- |
| Cortex | TIE1 | 1 | 1.26E-06 | 3.40E-05 | 1.04E-10 | 5.12E-11 | 1.40E-08 | 3.05E-09 |
|  | CDC20 | 1 | 1.10E-06 | 4.93E-03 | 3.57E-11 | 1.07E-10 | 5.12E-08 | 1.08E-04 |
|  | MPL | 1 | 1.68E-06 | 0.021 | 1.46E-08 | 5.65E-11 | 6.14E-09 | 5.71E-09 |
|  | IPO13 | 1 | 9.51E-09 | 1.56E-05 | 3.07E-08 | 8.15E-15 | 3.42E-13 | 4.09E-13 |
|  | PTPRF | 1 | 1.84E-08 | 1.42E-06 | 1.03E-11 | 6.65E-11 | 6.37E-14 | 3.92E-14 |
|  | MED8 | 1 | 6.27E-07 | 9.90E-11 | 7.85E-11 | 1.34E-11 | 4.75E-08 | 7.48E-08 |
|  | MEF2C | 5 | 2.59E-07 | 1.29E-03 | NA | 2.95E-07 | 1.62E-08 | 2.83E-12 |
|  | TMEM161B | 5 | 3.71E-07 | 9.87E-05 | 8.57E-08 | 3.37E-09 | 1.61E-09 | 4.20E-12 |
|  | XRN2 | 20 | 3.57E-08 | 7.58E-14 | 4.53E-10 | 9.26E-11 | 2.22E-10 | 2.02E-09 |
|  | KIZ | 20 | 8.48E-09 | 6.95E-15 | 2.97E-10 | 2.25E-10 | 3.73E-11 | 8.61E-14 |
|  | NKX2-4 | 20 | 3.50E-07 | 5.71E-06 | 2.70E-06 | 3.97E-12 | 3.50E-09 | 8.29E-11 |
| Basal Ganglia | MPL | 1 | 9.30E-08 | 0.676 | NA | 1.83E-12 | 1.45E-07 | 2.71E-08 |
|  | C1orf210 | 1 | 2.11E-07 | 1.18E-04 | NA | 1.97E-05 | 8.90E-12 | 3.18E-11 |
|  | MDFIC | 7 | 5.86E-07 | 2.67E-04 | NA | 4.23E-11 | 6.73E-10 | 4.36E-10 |
|  | NKX2-2 | 20 | 2.43E-08 | 1.51E-04 | NA | 2.28E-13 | 2.52E-10 | 2.21E-12 |
| Cerebellum | TIE1 | 1 | 4.67E-07 | 0.872 | 8.62E-12 | 4.48E-11 | 1.63E-10 | 7.60E-09 |
|  | MPL | 1 | 7.16E-07 | 4.97E-04 | NA | 1.92E-11 | 3.26E-10 | 1.34E-09 |
|  | ARTN | 1 | 1.15E-08 | 0.007 | 2.38E-13 | 1.32E-14 | 2.30E-07 | 8.70E-07 |
|  | IPO13 | 1 | 3.41E-08 | 0.637 | 5.85E-08 | 7.96E-14 | 6.37E-10 | 9.59E-10 |
|  | ATP6V0B | 1 | 5.28E-10 | 0.021 | 1.49E-14 | 5.37E-16 | 1.43E-16 | 1.50E-16 |
|  | ST3GAL3 | 1 | 4.95E-12 | 1.81E-10 | 6.09E-19 | 2.23E-20 | 1.09E-17 | 1.06E-12 |
|  | PTPRF | 1 | 2.77E-09 | 0.462 | NA | 1.35E-15 | 6.53E-09 | 9.85E-10 |
|  | CCDC24 | 1 | 7.10E-09 | 1.67E-04 | 3.66E-12 | 5.43E-15 | 2.77E-10 | 1.83E-11 |
|  | MED8 | 1 | 1.36E-07 | 4.59E-09 | 8.15E-10 | 8.46E-13 | 1.27E-09 | 4.17E-10 |
|  | FAM183A | 1 | 5.47E-10 | 0.847 | 0.208 | 6.81E-17 | 1.13E-09 | 1.12E-12 |
|  | SZT2 | 1 | 1.54E-06 | 1.10E-04 | 5.26E-11 | 5.72E-07 | 2.95E-07 | 4.16E-06 |
|  | HIGD1A | 3 | 1.40E-06 | 2.70E-05 | 0.116 | 0.103 | 0.005 | 4.51E-11 |
|  | HYAL3 | 3 | 1.55E-06 | 8.97E-08 | 6.44E-11 | 3.13E-10 | 6.97E-08 | 1.56E-07 |
|  | NKX2-2 | 20 | 1.21E-06 | 0.605 | 1.09E-10 | 5.14E-11 | 3.41E-07 | 2.01E-06 |

### Fine-Mapping

We conducted fine mapping among genomic regions with multiple risk genes (Table 2). This included three regions in the cortex, one region in the basal ganglia, and one region in the cerebellum. Genes that were the sole significant signal in a region were considered the best-case gene. Of the 29 identified genes across tissues, 23 genes were found to be putatively causal signals with GIFT after FDR correction, with 11 risk genes in the cortex (chr1:42800982-44468022; chr5:87689633-89404257; chr20:20625983-21897526), two in basal ganglia (chr1:42781877-43852772), and 10 in cerebellum (chr1:42645153-44496528) (Figures S7-S12). Five genes were the only identified genes in their regions and were therefore considered independent signals. One gene (ATP6V0B) in the cerebellum, region chr1:42645153-44496528, was not statistically significant after fine-mapping.

**Table 2:** GIFT fine-mapping results for regions containing multiple ADHD TWAS loci. . For each tissue and genomic region, fine-mapping *P* values are reported for the omnibus OTTERS and for individual prediction methods. Multiple testing correction using the false discovery rate (FDR) was applied to the OTTERS fine-mapping *P* values. Missing values, noted by NA, indicate that a given

| Tissue | Gene | Region | lassosum | P0.001 | P0.05 | PRScs | SDPR | OTTERS |
| --- | --- | --- | --- | --- | --- | --- | --- | --- |
| **Cortex** | TIE1 | chr1:42800982-44468022 | 1.11E-11 | 0.002 | 2.83E-11 | 2.39E-11 | 0.003 | 5.48E-11 |
|  | CDC20 | chr1:42800982-44468022 | 0.349 | 5.73E-04 | 0.045 | 0.060 | 0.341 | 0.003 |
|  | MPL | chr1:42800982-44468022 | 5.14E-27 | 0.495 | 0.193 | 3.00E-08 | 1.95E-22 | 5.65E-26 |
|  | IPO13 | chr1:42800982-44468022 | 0.013 | 1.68E-05 | 0.014 | 3.52E-09 | 1.38E-08 | 1.93E-08 |
|  | PTPRF | chr1:42800982-44468022 | 0.042 | 3.43E-53 | 2.08E-44 | 1.43E-16 | 1.60E-13 | 1.89E-51 |
|  | MED8 | chr1:42800982-44468022 | 0.012 | 0.190 | 0.433 | 0.197 | 0.012 | 0.028 |
|  | MEF2C | chr5:87689633-89404257 | 0.138 | 2.33E-24 | 7.3E-17 | 7.66E-16 | 4.18E-30 | 5.75E-29 |
|  | TMEM161B | chr5:87689633-89404257 | 2.41E-21 | 1.17E-20 | 8.0E-34 | 5.56E-08 | 0.003 | 1.47E-32 |
|  | XRN2 | chr20:20625983-21897526 | 6.15E-10 | 0.328 | 0.233 | 1.15E-04 | 0.008 | 4.83E-09 |
|  | KIZ | chr20:20625983-21897526 | 0.013 | 0.151 | 0.016 | 7.76E-05 | 1.43E-08 | 8.71E-08 |
|  | NKX2-4 | chr20:20625983-21897526 | 1.77E-49 | 2.97E-22 | 1.54E-28 | 1.52E-06 | 6.35E-09 | 4.86E-48 |
| **Basal Ganglia** | MPL | chr1:42781877-43852772 | 0.028 | NA | 1.89E-09 | 5.01E-18 | 7.47E-16 | 3.98E-17 |
|  | C1orf210 | chr1:42781877-43852772 | 2.10E-09 | NA | 4.75E-11 | 3.22E-16 | 4.96E-13 | 1.29E-15 |
| **Cerebellum** | TIE1 | chr1:42645153-44496528 | 0.003 | 0.684 | 3.03E-11 | 1.69E-18 | 8.11E-13 | 1.85E-17 |
|  | MPL | chr1:42645153-44496528 | 1.58E-11 | NA | 3.91E-21 | 1.16E-06 | 0.997 | 8.61E-20 |
|  | ARTN | chr1:42645153-44496528 | 0.8577 | 4.42E-14 | 6.45E-44 | 4.77E-07 | 0.007 | 2.84E-42 |
|  | IPO13 | chr1:42645153-44496528 | 0.7501 | 0.928 | 6.48E-04 | 0.008 | 9.73E-04 | 0.002 |
|  | ATP6V0B | chr1:42645153-44496528 | 0.0351 | 0.054 | 0.907 | 0.373 | 0.018 | 0.053 |
|  | ST3GAL3 | chr1:42645153-44496528 | 3.62E-04 | 1.68E-06 | 0.313 | 0.004 | 0.043 | 0.002 |
|  | PTPRF | chr1:42645153-44496528 | 0.006 | NA | 0.104 | 0.527 | 0.150 | 0.026 |
|  | CCDC24 | chr1:42645153-44496528 | 2.69E-20 | 0.462 | 0.385 | 8.25E-16 | 8.43E-19 | 3.83E-19 |
|  | MED8 | chr1:42645153-44496528 | 0.038 | 9.29E-06 | 2.91E-05 | 0.930 | 0.097 | 2.13E-04 |
|  | FAM183A | chr1:42645153-44496528 | 0.521 | 0.003 | 4.51E-06 | 0.111 | 0.038 | 3.97E-05 |
|  | SZT2 | chr1:42645153-44496528 | 0.016 | 0.265 | 0.019 | 0.709 | 0.562 | 0.040 |

method did not yield a valid prediction model for that gene-tissue pair in the TWAS

### Colocalization

We identified several genes with strong evidence that the ADHD GWAS and eQTL signals share a causal variant (PP.H4 > 0.7) (Table 3). In the cortex, fives genes presented with evidence of strong colocalization: *TIE1* (PP.H4=0.9), *PTPRF* (PP.H4=0.99), *KIZ* (PP.H4=0.99), *MED8* (PP.H4=0.73), and *XRN2* (PP.H4=0.72) (Table 3; Table S5). In the cerebellum the *HYAL3* gene presented with strong evidence for colocalization across ten independent credible sets, with PP.H4 values between 0.99 and 1.0 (Table 3; Table S6). No basal ganglia genes exceeded the PP.H4 threshold to declare colocalization.

**Table 3:** Colocalization results for ADHD TWAS loci. Bayesian colocalization analyses assessing whether ADHD GWAS and eQTL association signals share a common causal variant across tissues. Tissue gene-pairs with detectable eQTL signals and sufficient SNP overlap were evaluated. For each tissue-gene pair, the number of SNPs tested, the number of colocalizing credible set (CS) pairs, the maximum posterior probability of a shared causal variant (PP.H4), and the lead GWAS and eQTL variants are reported. Higher PP.H4 values indicate stronger evidence of colocalization. Evidence for strong colocalization is defined as PP.H4 > 0.8 and moderate colocalization as 0.5> PP.H4 < 0.8.

| Tissue | Gene Symbol | Gene chr | Gene position | Number of SNPs | Colocalizing CS pairs | Maximum PP.H4 | Lead GWAS variant | Lead eQTL variant |
| --- | --- | --- | --- | --- | --- | --- | --- | --- |
| Cortex | PTPRF | 1 | 43525187 | 1237 | 1 | 0.999997911 | 1:43939667 | 1:43939667 |
|  | TIE1 | 1 | 43300982 | 1334 | 1 | 0.999997494 | 1:43541977 | 1:43541977 |
|  | KIZ | 20 | 21125983 | 1090 | 1 | 0.999975397 | 20:21194910 | 20:21194910 |
|  | MED8 | 1 | 43389808 | 1351 | 1 | 0.724793426 | 1:43549154 | 1:43541977 |
|  | XRN2 | 20 | 21303331 | 1108 | 1 | 0.718344018 | 20:21194910 | 20:21234363 |
|  | CDC20 | 1 | 43358981 | 1357 | 0 | 0.417908926 | 1:43548609 | 1:43547684 |
|  | IPO13 | 1 | 43946950 | 1236 | 0 | 0.297830409 | 1:43939667 | 1:43944271 |
|  | TMEM161B | 5 | 88269476 | 965 | 0 | 0.023452263 | 5:88458988 | 5:88091755 |
|  | MPL | 1 | 43337849 | 1349 | 0 | 0.000562438 | 1:43541977 | 1:43407812 |
| Cerebellum | HYAL3 | 3 | 50299405 | 822 | 10 | 1 | 3:50198940 | 3:50198940 |

### Functional Enrichment

In the gene ontology over-enrichment analysis, we identified statistically significantly (FDR<0.05) enriched genes in the cortex and basal ganglia ,but not in the cerebellum (Tables S2-S4). Across 62 terms in the basal ganglia (58 Biological Process, 4 Molecular Function), *NKX2-2* was enriched for neurodevelopmental pathways such as neuron fate determination, ventral spinal cord interneuron differentiation, and oligodendrocyte development. *MPL* mapped to platelet hematopoietic/immune biology, and included terms such as platelet formation and morphogenesis, neutrophil/leukocyte homeostasis, cytokine signaling/receptor activity. *MDFIC* was enriched in transcriptional/nuclear transport. Overlap across genes were minimal apart from one term “positive regulation of cell development” shared by *NKX2-2* and *MPL*. In the cortex, the two significant terms were related to platelet morphogenesis and platelet formation shared by *MPL*/*MDFIC*. Using the STRING database, the PPI p-value was 1.12E-04, indicating that the gene network had significantly more interactions than expected and that the proteins are at least partially biologically connected. The network was significantly enriched in ADHD and autism spectrum disorder-related reference publications (Figures S13-S14).

## Discussion

In this multi-tissue TWAS of ADHD, we utilized the OTTERS framework, which aggregates five PRS-based GReX models to conduct a two-stage TWAS using summary-level eQTL and GWAS data. We identified 29 risk genes whose genetically regulated expressions were significantly associated with ADHD risk across the cortex (11 genes), basal ganglia (4 genes), and cerebellum (14 genes). Among regions with multiple risk genes, fine-mapping with GIFT prioritized 23 of these genes as putatively causal and six genes exhibited moderate or strong evidence of colocalization.

Several of the genes identified in our TWAS have been previously implicated in ADHD, identified through approaches including GWAS, TWAS, and gene-based analysis. In the cortex, we replicated associations for 10 previously reported ADHD genes (*TIE1* (10,31,32), *CDC20* (31), *IPO13* (31), *PTPRF* (3,4,31,32), *MED8* (4,10,31), *MEF2C* (10,31), *TMEM161B* (3), *XRN2* (4), *KIZ* (4), and NKX2-4 (4)), while *MPL* is a novel finding. In the cerebellum we replicated an additional six previously reported (*ARTN* (10,31), *ATP6V0B* (31), *ST3GAL3* (3,31,32), *CCDC24* (10,31), *SZT2* (31), and *HYAL3* (4), and identified three novel associations (*FAM183A*, *HIGD1A*, *NKX2*-2). In the basal ganglia all four associated genes are novel (*MPL*, *C1orf210*, *MDFIC*, *NKX2-2*).

Compared with the 2023 ADHD GWAS meta-analysis which our study is based on, four genes overlapped (*PTPRF*, *XRN2*, *NKX2-4*, *ST3GAL3*) with their SNP-level findings (4). We also observed variation in tissue-specificity relative to previous TWAS. For example, in the 2023 TWAS conducted in dorsolateral prefrontal cortex, *MED8* and *KIZ* overlapped with our cortex results, while *MED8* and *HYAL3* overlapped with our cerebellum results (4). In a 2019 TWAS based on 2019 GWAS summary statistics (3), *ARTN*, *CCDC24*, and *MED8* identified in cerebellum overlapped with our cerebellum findings, while TIE1 overlapped with our cortex findings (10). Notably, we detected *MED8* and *TIE1* in both cortex and cerebellum. *TIE1*, *PTPRF*, and *MED8* exhibited the most robust result, identified in at least three previous studies.

We identified 5 novel risk genes, including *MPL*, *C1orf210*, *MDFIC, FAM183A*, *HIGD1A*, and *NKX2-2*. *MPL* is a novel risk gene that was associated with all three tissues and was functionally enriched in the basal ganglia and cortex. It primarily plays a role in making the thrombopoietin receptor protein, which promotes the growth and division of cells, in particular the production of platelets, erythrocytes, and megakaryocytes. It has been previously shown that the *MPL* gene is expressed in neurons of the central nervous system (33,34). While traditionally associated with blood cell disorder, some studies have investigated the role of MPL in neurodevelopment. For example, one previous study identified an association between increased platelet count among patients with ASD, suggesting that genetic factors associated with platelet counts may be associated, but not fully penetrant for ASD (35). One gene-based analysis of potential gene drug targets for ADHD identified *MPL* as a potential druggable gene(36). Further studies should be conducted to investigate the relationship between platelet function and child psychopathologies. *NKX2-2* was identified as a risk gene in the basal ganglia and cerebellum and plays a role in cell differentiation and development of the nervous system, including neuroendocrine and glial differentiation and oligodendrocyte differentiation. It was also significantly overrepresented for these factors. *NKX2-2* is expressed across all brain regions, most highly in white-matter, which is commonly altered in ADHD (33,37). It has previously been identified as a significant risk gene in autism spectrum disorder (38,39). More research is needed to determine the role of *NKX2-2* in the etiology of ADHD and related disorders. *MDFIC* is a transcriptional modular involved in transcriptional regulation and was identified as a novel risk gene in the basal ganglia. It is highly expressed across multiple brain regions (36). While not previously associated with child psychopathologies, it has been implicated as a pleiotropic locus between anti-social behavior and ADHD (40).

Notably, we identified four genes exhibiting strong colocalization, indicating that ADHD risk and regulatory variants influencing expression of these genes likely represent shared causal signals. In the cortex, *TIE1*, *PTPRF*, and *KIZ* exhibited strong colocalization, while in the cerebellum *HYAL3* exhibited strong evidence of colocalization and was the only sole significant signal in the gene region. No genes colocalized in the basal ganglia, likely due to the small sample size and limited statistical power of the eQTL summary statistics. *TIE1* is expressed across several tissues of the brain, particularly in vascular endothelial cells (33). It’s primarily involved in angiogenesis and blood vessel development (41). *TIE1* dysfunction can lead to vascular destabilization and activation of inflammatory pathways that influence neuronal development (42,43). *PTPRF* is a cell adhesion receptor involved in regulation of cellular processes such as cell growth and differentiation and may play a potential role in axon formation and synaptogenesis during neurodevelopment (44–46). It is expressed in astrocytes across several brain tissues (33). *KIZ* is a centrosomal protein that regulates spindle formation during mitosis (33). It has been implicated as a significant risk gene of both ADHD and ASD (4,39,47). *HYAL3* gene plays a role in degradation of hyaluronic acid, which is a major component of the extracellular matrix (33). It has previously been associated with inflammatory cell type morphology and identified as an ADHD risk gene in cortex tissue (4,48). Identification in the cerebellum is a novel finding.

Interestingly, a few of our associated genes also overlapped with previous findings in Autism Spectrum Disorder (ASD), suggesting potential pleiotropy. *NKX2-4*, *KIZ*, *XRN2*, and *NKX2-2*, were previously identified in a gene-based analysis of ASD(47), while genes, *MEF2C*, *TMEM161B*, *XRN2*, *KIZ*, *NKX2-4*, and *NKX2-2* were identified in a multi-variable regression analysis of both ADHD and ASD (39). The 2023 ADHD GWAS conducted an LD score regression to assess the genetic overlap between ADHD and ASD and identified a significant genetic correlation (genome-wide r=0.42; standard error=0.05). They also found that 84% of concordant and discordant ADHD risk variants also influenced ASD. Together this indicates that many genetic variants contributing to ADHD risk also influences ASD risk through overlapping biological mechanisms.

We observed both cross-tissue overlap and tissue-specific signals. Four genes (*TIE1*, *IPO13*, *PTPRF*, and *MED8*) were identified in both cortex and cerebellum. *NKX2-2* was identified in the basal ganglia and cerebellum. *MPL* was detected in all three tissues. This suggests that many genes are co-expressed and that region-wide dysregulation in gene expression likely plays a role in the etiology of ADHD. On the other hand, several genes were tissue-specific, including 6 of 10 genes in the cortex, 2 of 4 genes in the basal ganglia, and 8 of 14 genes in the cerebellum. Tissue-specific findings may in-part reflect differences in eQTL reference panel sample sizes which can influence the statistical power to detect cross-tissue associations (14). Additionally, as many genes share transcriptional regulation across tissues, cross-tissue eQTL effects can lead to correlated TWAS signals, potentially obscuring true tissue-specific mechanisms (49). Assessing only the most mechanistically relevant tissue to ADHD, as we have here, should mitigate tissue bias while integrating fine-mapping and colocalization further ensures the robustness of our findings and prioritizes genes with shared causal genetic variants(50). Nevertheless, these results highlight the importance of both cross-tissue replication and multi-tissue analyses for uncovering region-specific associations.

Several genes identified in this study are not directly involved in neurotransmission or neurodevelopment, but rather implicate vascular, immune, and other systemic biological processes. Recent studies have suggested that brain vascularization and in particular endothelial cells, supports neuronal development including, neurogenesis and neuronal migration, and may contribute to risk for neurodevelopmental disorders (51). Additionally, several studies have linked elevated inflammatory proteins during pregnancy to increased risk of ADHD and related disorders in offspring (52).

Strengths of this study include the use of OTTERS to aggregate five PRS-based GReX training approaches with summary-level reference data, which helps to increase power and confidence in results. Additionally, this study was conducted across three biologically relevant brain tissue, which helps to mitigate tissue bias and builds upon previous TWAS conducted in the cortex (4,7). Limitations of this study include the limited sample size of the eQTL reference dataset, which may have reduced our power to detect true signals. Additionally, this study was conducted among participants from European ancestry, which restricts generalizability to the European population. This emphasizes the need for future work to validate these findings in diverse ancestries. Additionally, eQTL from bulk tissue results in uneven sample sizes across regions, which can reduce power and lead to false negatives (53,54). Future work using single-cell data and larger eQTL reference panels is necessary for further replication (55).

In conclusion, we identified 29 ADHD-associated genes across cortex, basal ganglia, and cerebellum, including six genes novel to ADHD. Concordance with prior GWAS/TWAS and gene-based studies supports the robustness of several signals, while fine-mapping and colocalization prioritized several candidates for mechanistic follow-up. Identifying differentially expressed ADHD risk genes provides novel insights for potential targets in translational research and drug discovery.

## Data Availability

All data used in this study are publicly available online: GWAS summary statistics are located at:https://www.med.unc.edu/pgc/download-results/
eQTL summary statistics are located at: https://www.metabrain.nl
TWAS summary statistics are available upon reasonable request to the authors.

https://www.metabrain.nl

## Acknowledgements

This work was supported by National Institutes of Health grant awards RF1AG071170 (QD, MPE), R35GM138313 (QD, JY), R01AG089703 (JY), U01AG088425 (AH). SA was supported by the National Institute of Environmental Health Sciences (T32ES012870) and (F31ES037540). The content is solely the responsibility of the authors and does not necessarily represent the official views of the National Institutes of Health.

We would like to thank all research participants for making this work possible. The following are web resources for this study: GWAS summary statistics: https://www.med.unc.edu/pgc/download-results/; eQTL summary statistics: https://www.metabrain.nl; OTTERS R package: https://github.com/daiqile96/OTTERS; coloc R package: https://chr1swallace.github.io/coloc/index.html GIFT R package: https://github.com/yuanzhongshang/GIFT ; clusterProfiler R package: https://bioconductor.org/packages/release/bioc/html/clusterProfiler.html

