## Supplemental Figures for "Multi-tissue transcriptome-wide association study identifies 29 risk genes associated with attention-deficit/hyperactivity disorder"

615 Michael Street

Suite 305L

Atlanta, GA 30322

404-712-8289

**Figure S1:** Cortex TWAS QQ plot for each method before genomic control. Lambda values are 1.55,1.65,1.61,1.66,1.71,1.74, respectively.

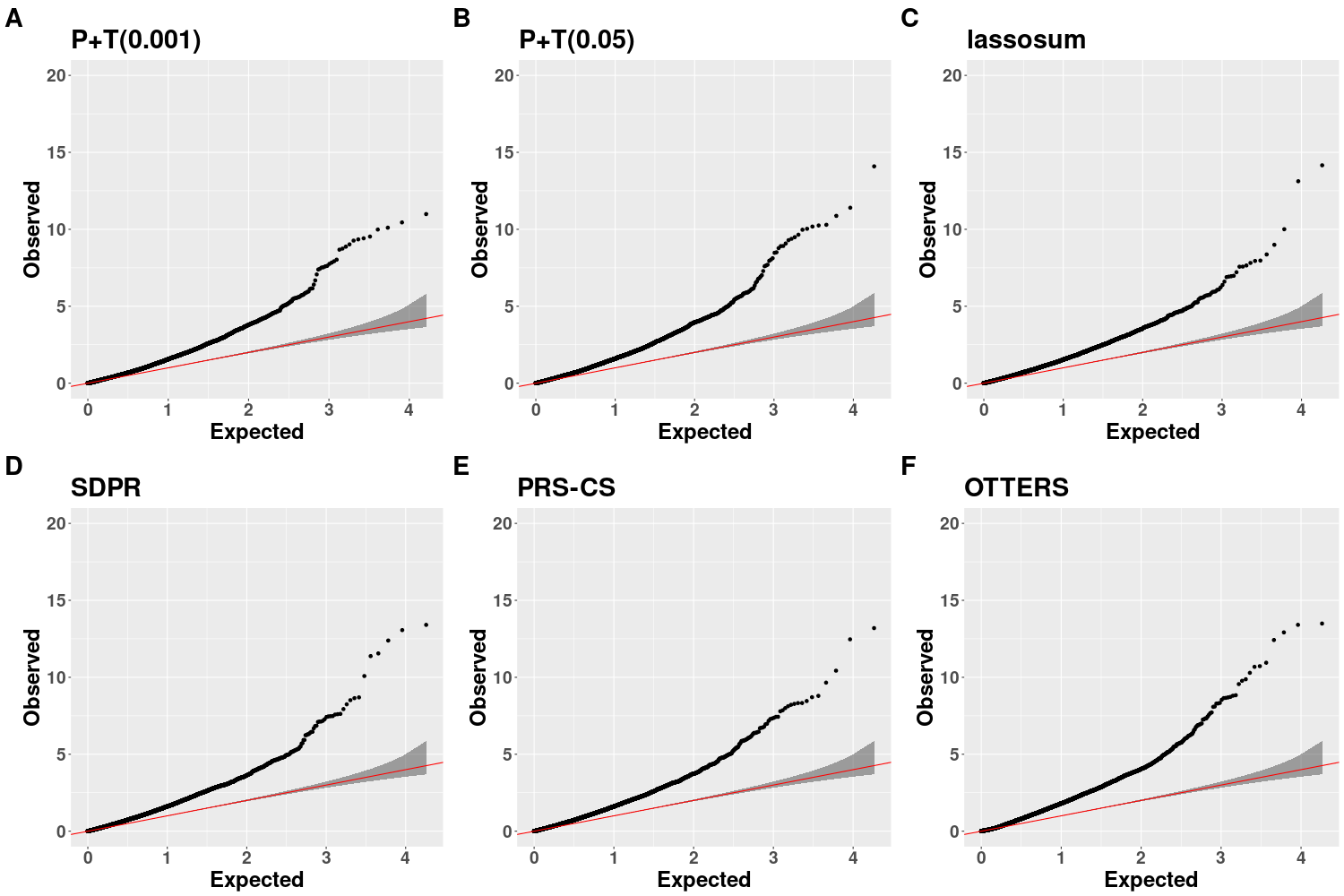

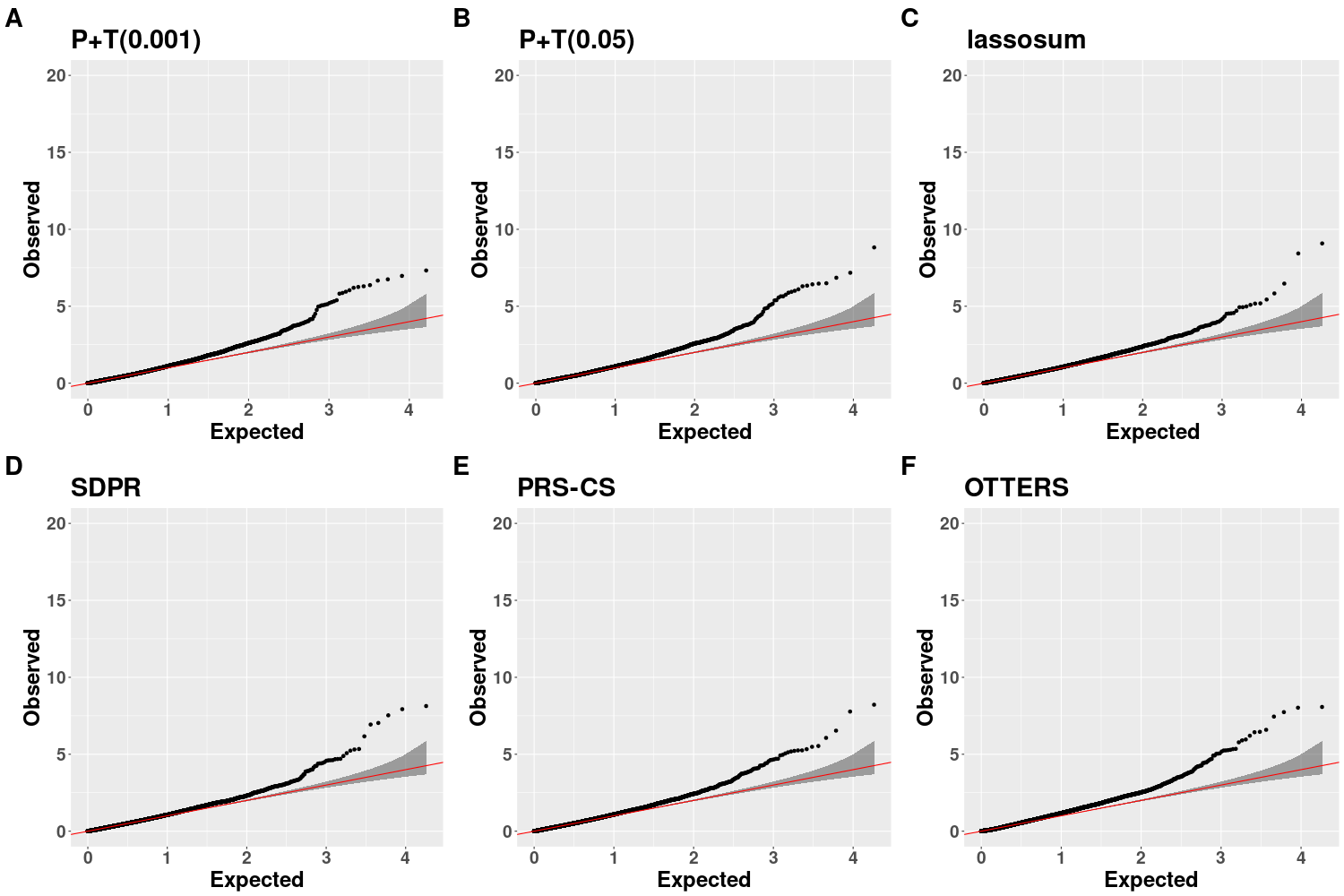
**Figure S2:** Cortex TWAS QQ plot for each method after genomic control. Lambda values are all 1.

**Figure S3:** Cerebellum TWAS QQ plots for all methods before genomic control. Lambda values are 1.48, 1.60, 1.53, 1.69, 1.71, 1.73, respectively.

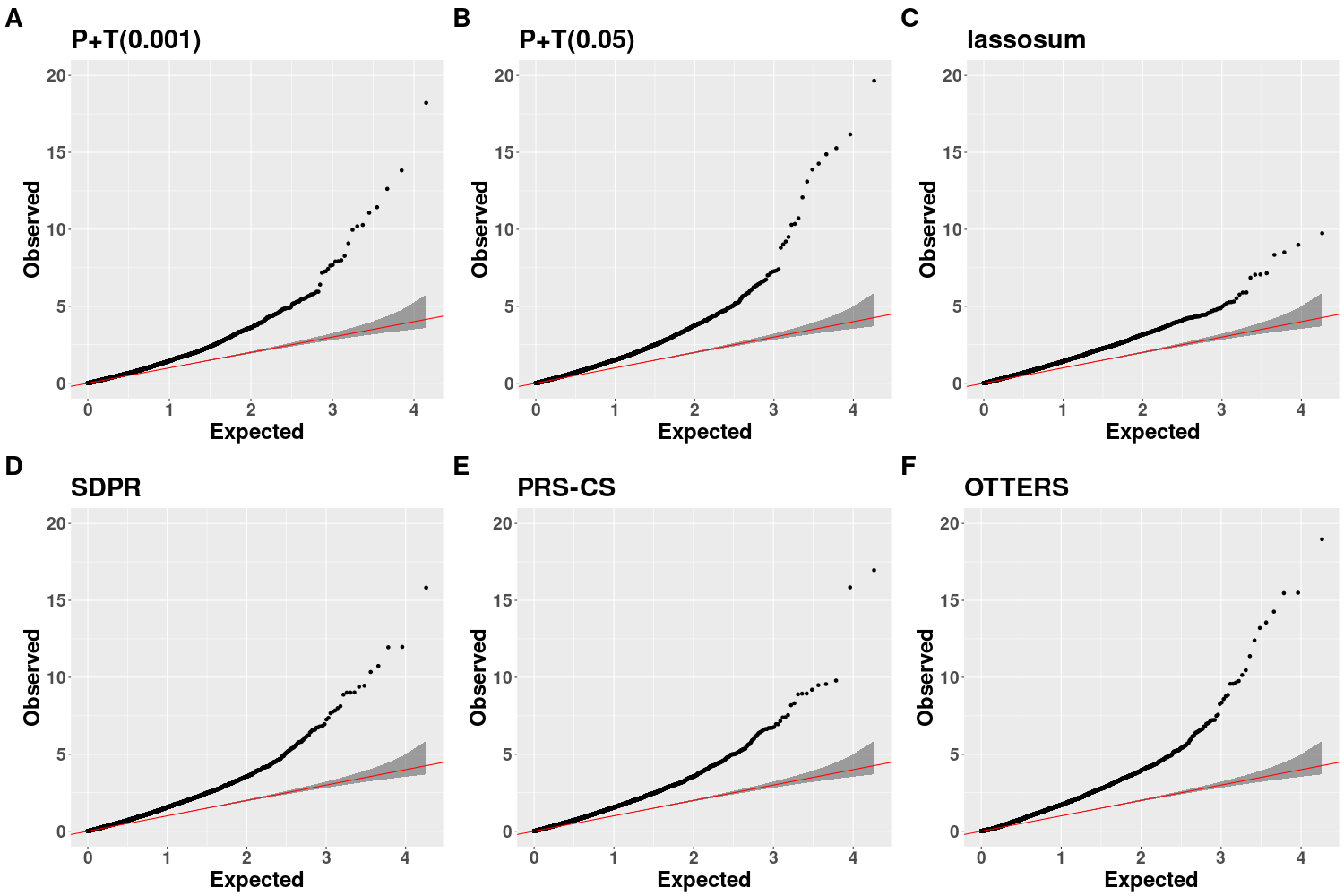

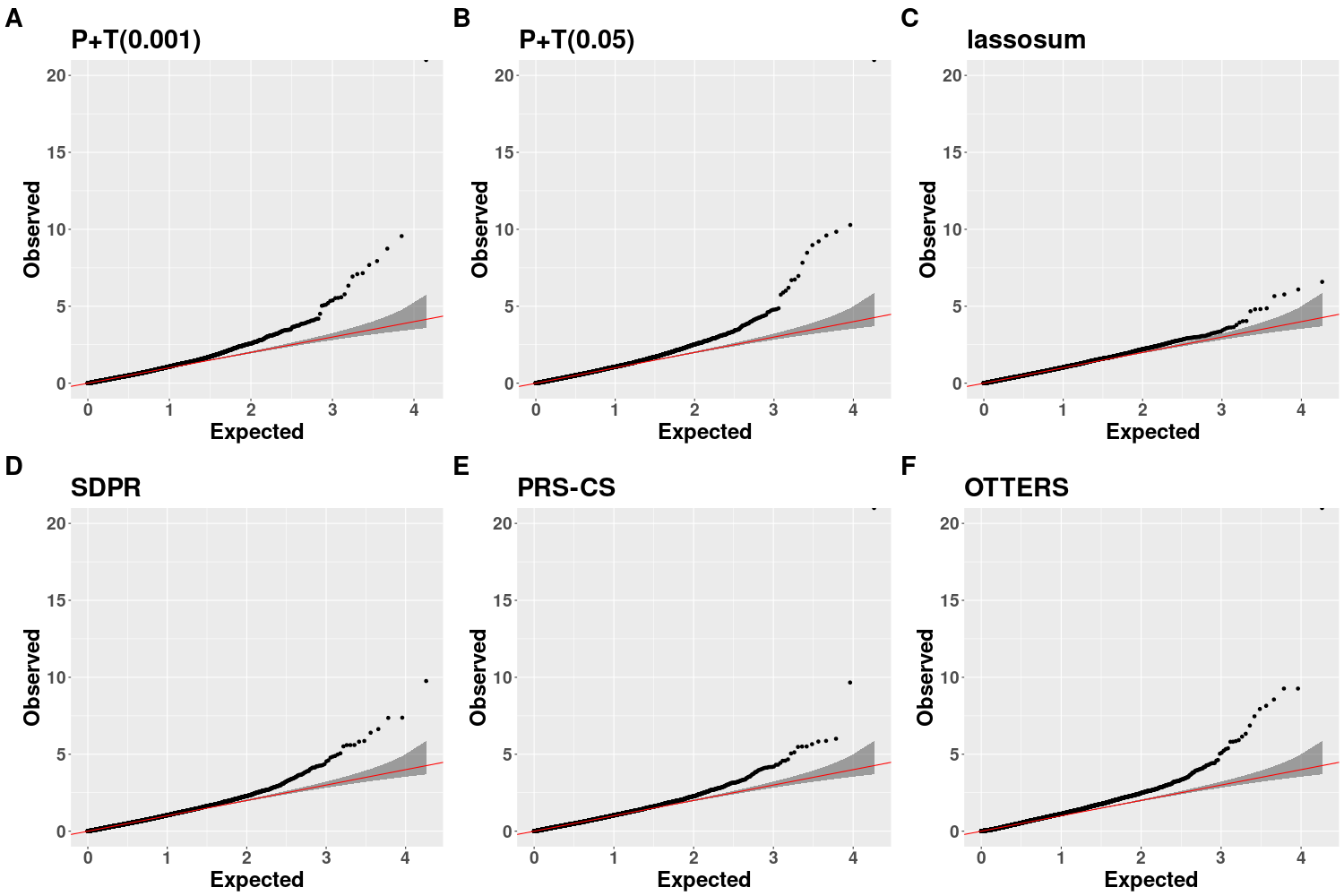
**Figure S4:** Cerebellum TWAS QQ plots for all methods after genomic control. All lambda values are 1.

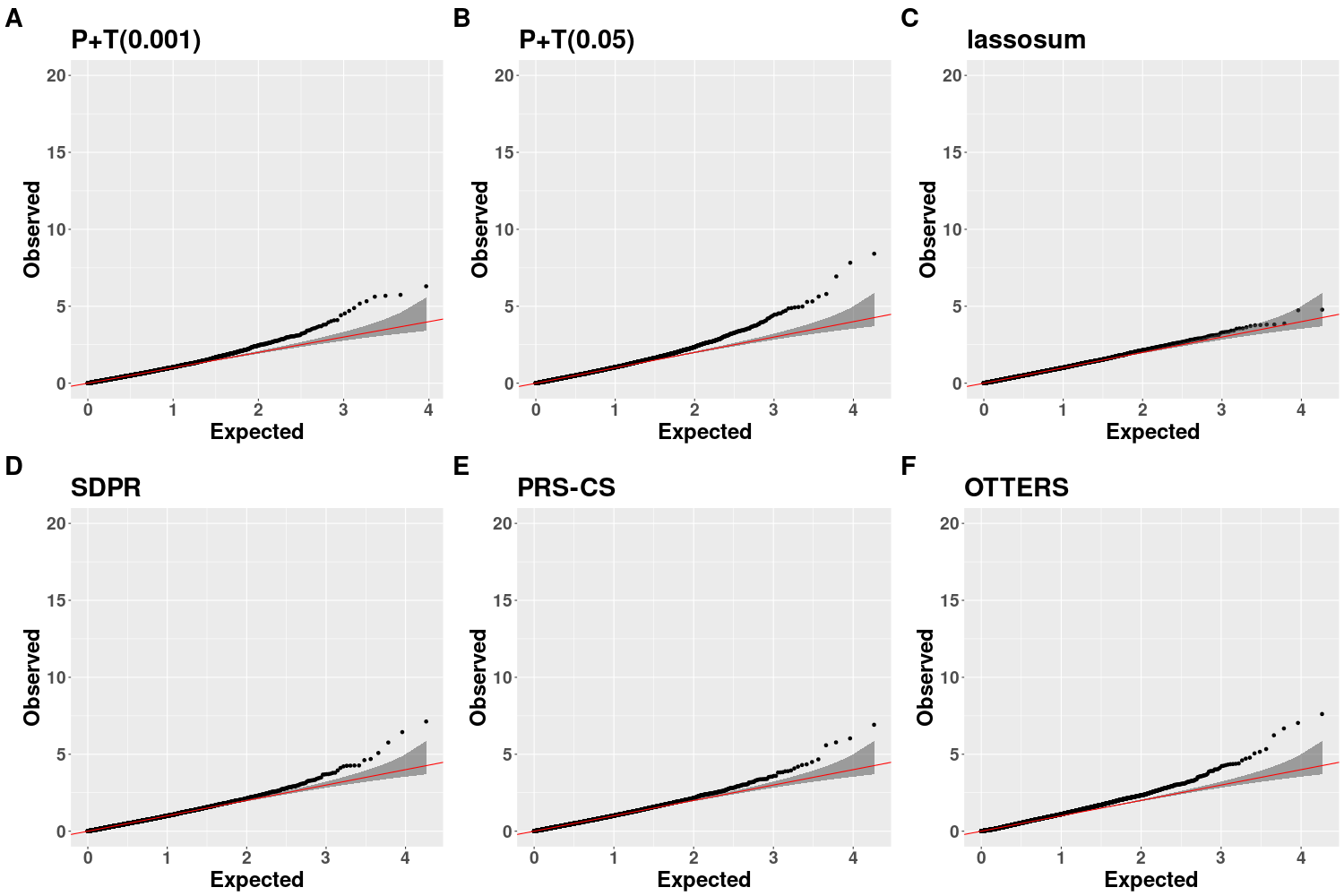
**Figure S5:** Basal Ganglia TWAS QQ plots for all methods before genomic control. Lambda values are 1.44, 1.55, 1.47, 1.70, 1.66, 1.65, respectively.

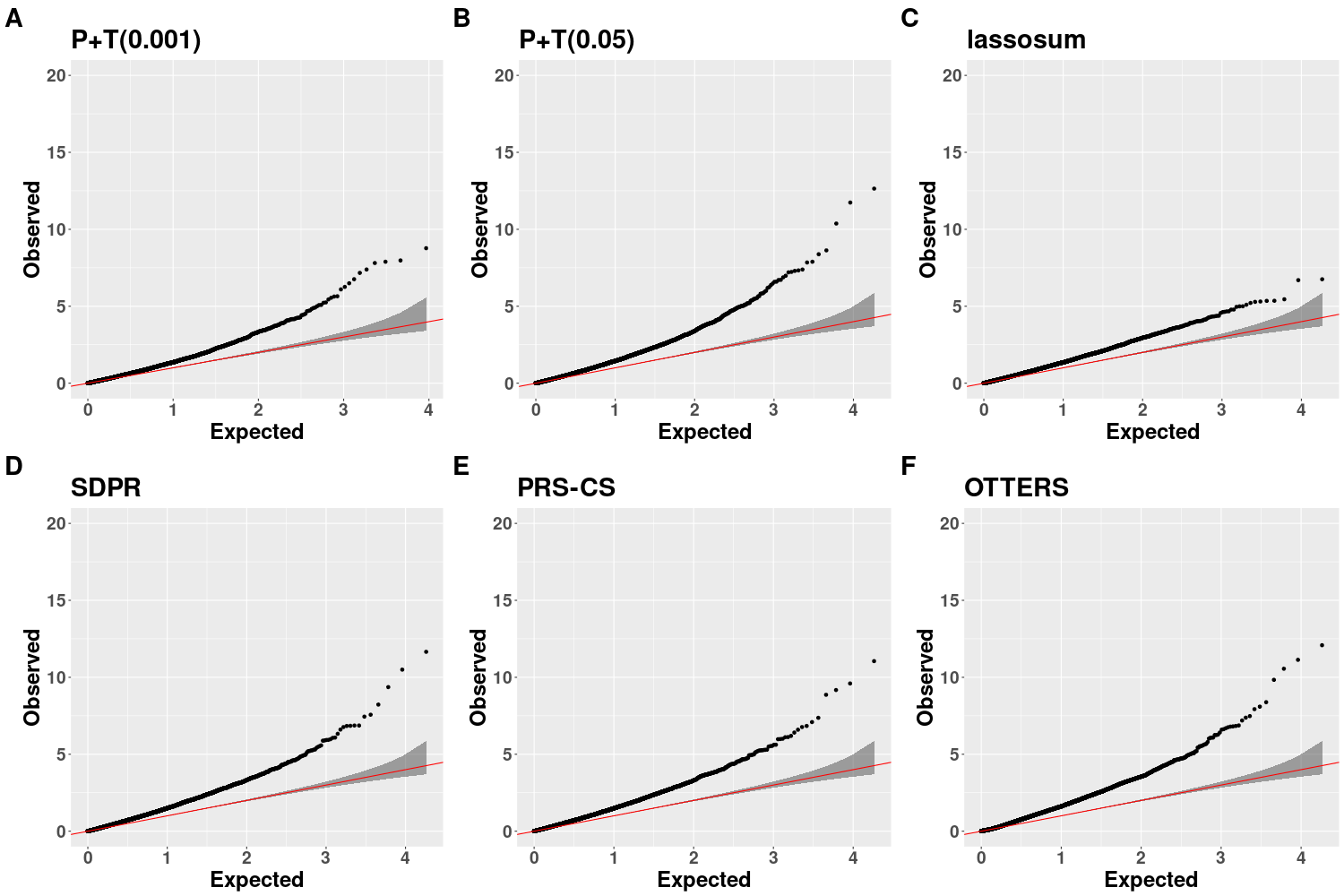
**Figure S6:** Basal Ganglia TWAS QQ plots for all methods after genomic control. All lambda values are 1.

**Figure S7: Overlap of significant ADHD TWAS genes that showed evidence of fine-mapping across brain tissues.** A) Venn diagram showing overlap of TWAS-significant genes that showed evidence of fine-mapping across cortex, basal ganglia, and cerebellum. B) UpSet plot summarizing the number of shared and tissue-specific TWAS-significant genes that showed evidence of fine-mapping across tissues.

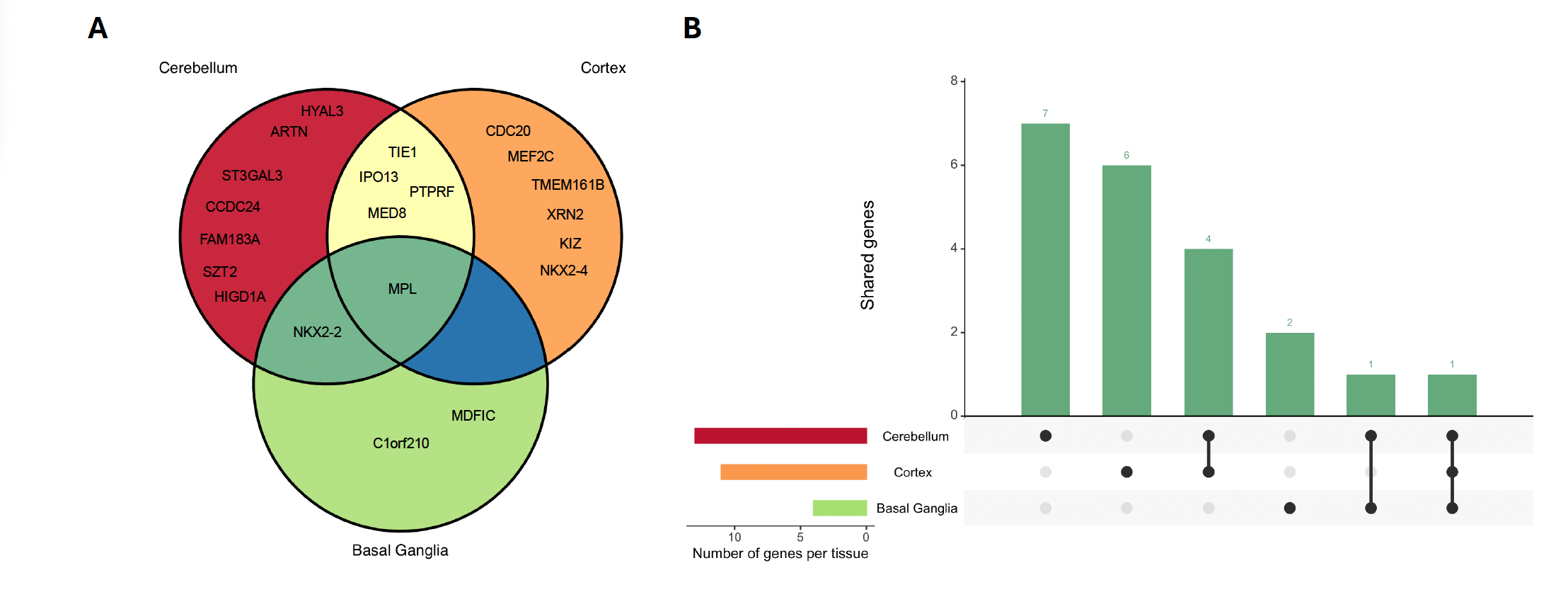

**Figure S8: GIFT results for cortex at region chr1:42800982-44468022.** The gray circle represents the -log10(p-value) of each SNP from the GWAS, the blue square represents the -log10(p-value) of each gene from the OTTERS TWAS, and the red diamond represents the -log10(p-value) of each gene from GIFT. The red dashed line represents the GIFT significance threshold of 0.05. All genes that were statistically significant in the TWAS are labeled.

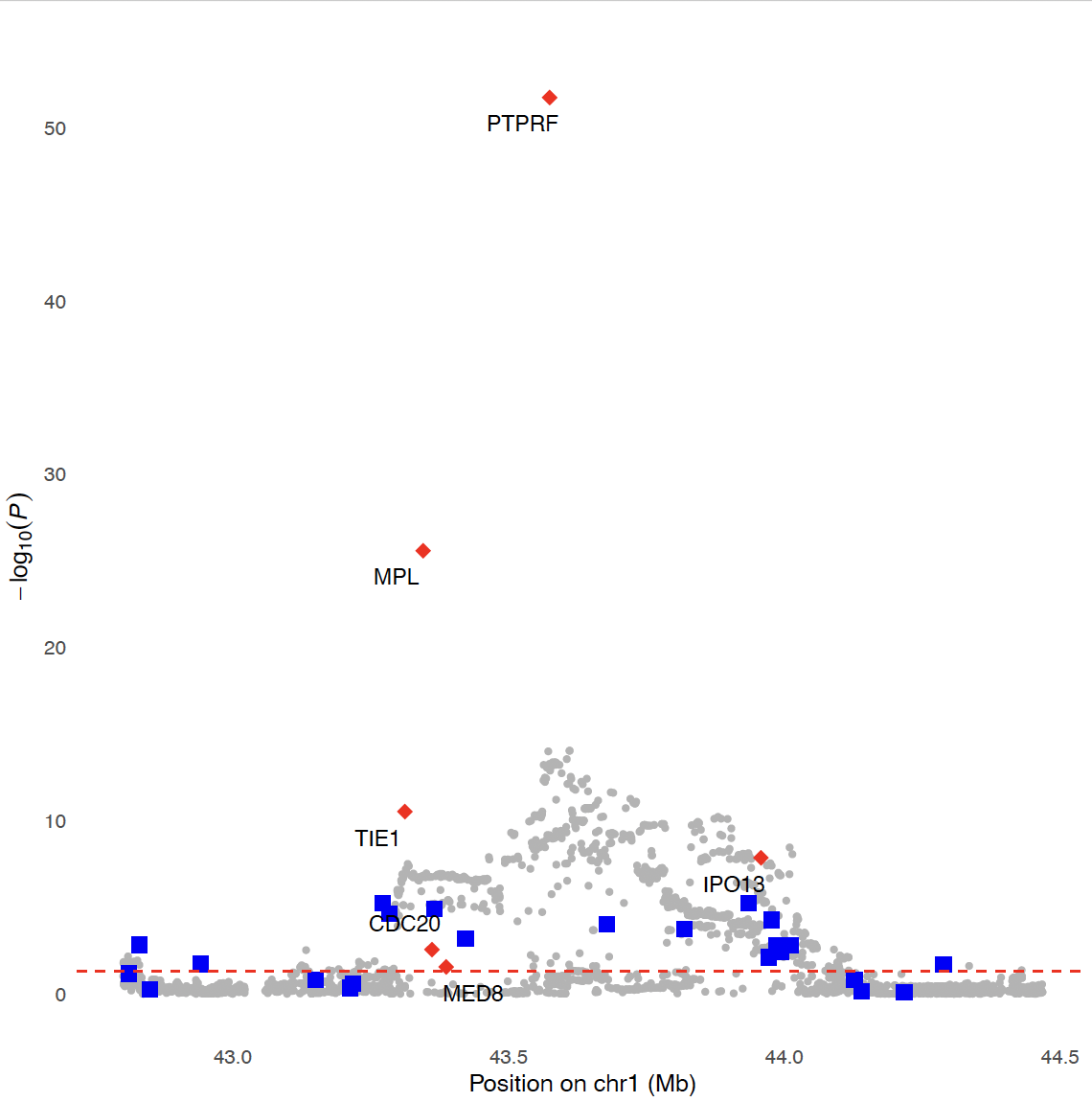

**Figure S9:** **GIFT results for cortex at region chr5:87689633-89404257.** The gray circle represents the -log10(p-value) of each SNP from the GWAS, the blue square represents the -log10(p-value) of each gene from the OTTERS TWAS, and the red diamond represents the -log10(p-value) of each gene from GIFT. The red dashed line represents the GIFT significance threshold of 0.05. All genes that were statistically significant in the TWAS are labeled. In this region, there were only two genes in the TWAS.

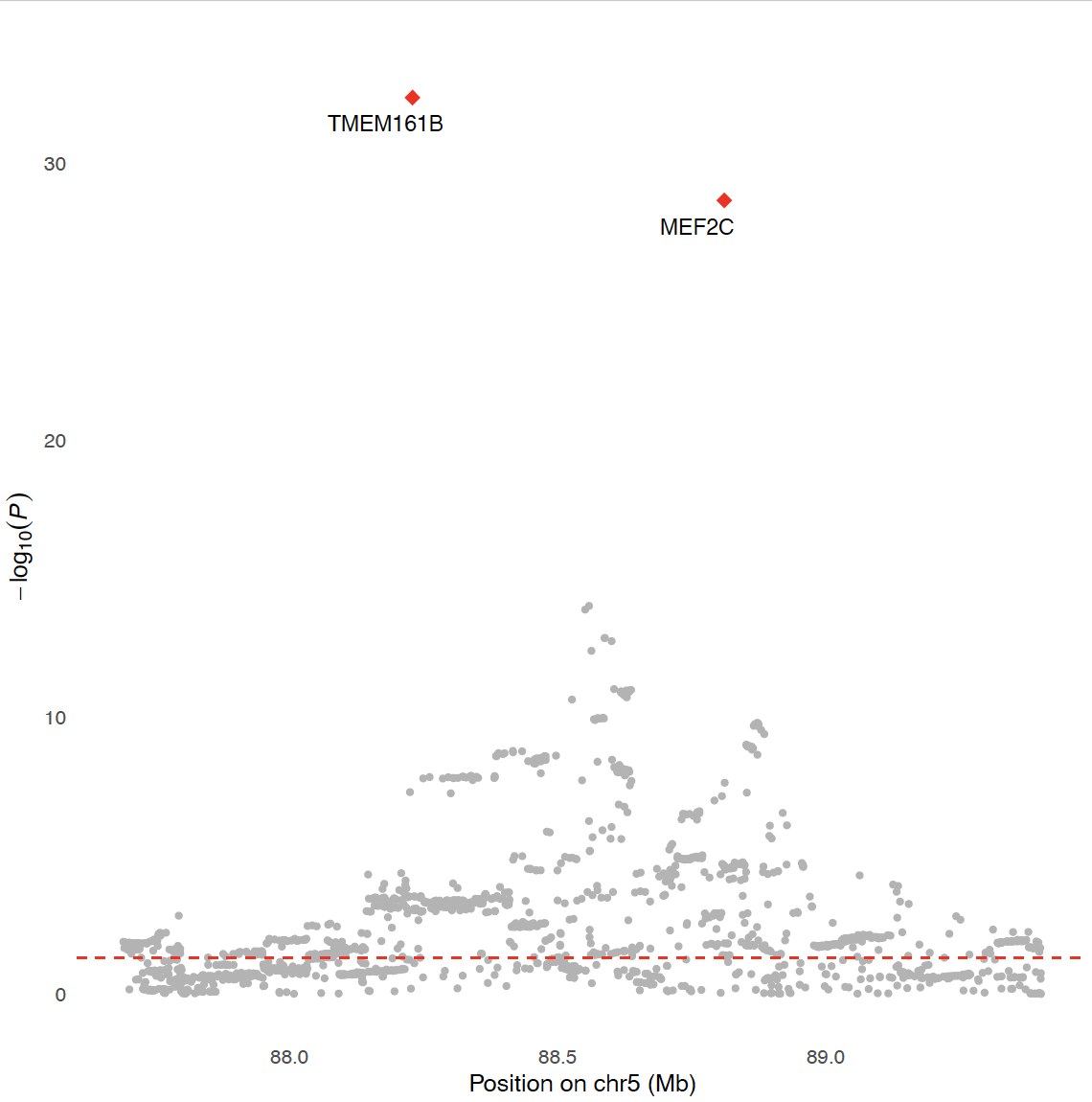

**Figure S10:** **GIFT results for cortex at region chr20:20625983-21897526.** The gray circle represents the -log10(p-value) of each SNP from the GWAS, the blue square represents the -log10(p-value) of each gene from the OTTERS TWAS, and the red diamond represents the -log10(p-value) of each gene from GIFT. The red dashed line represents the GIFT significance threshold of 0.05. All genes that were statistically significant in the TWAS are labeled.

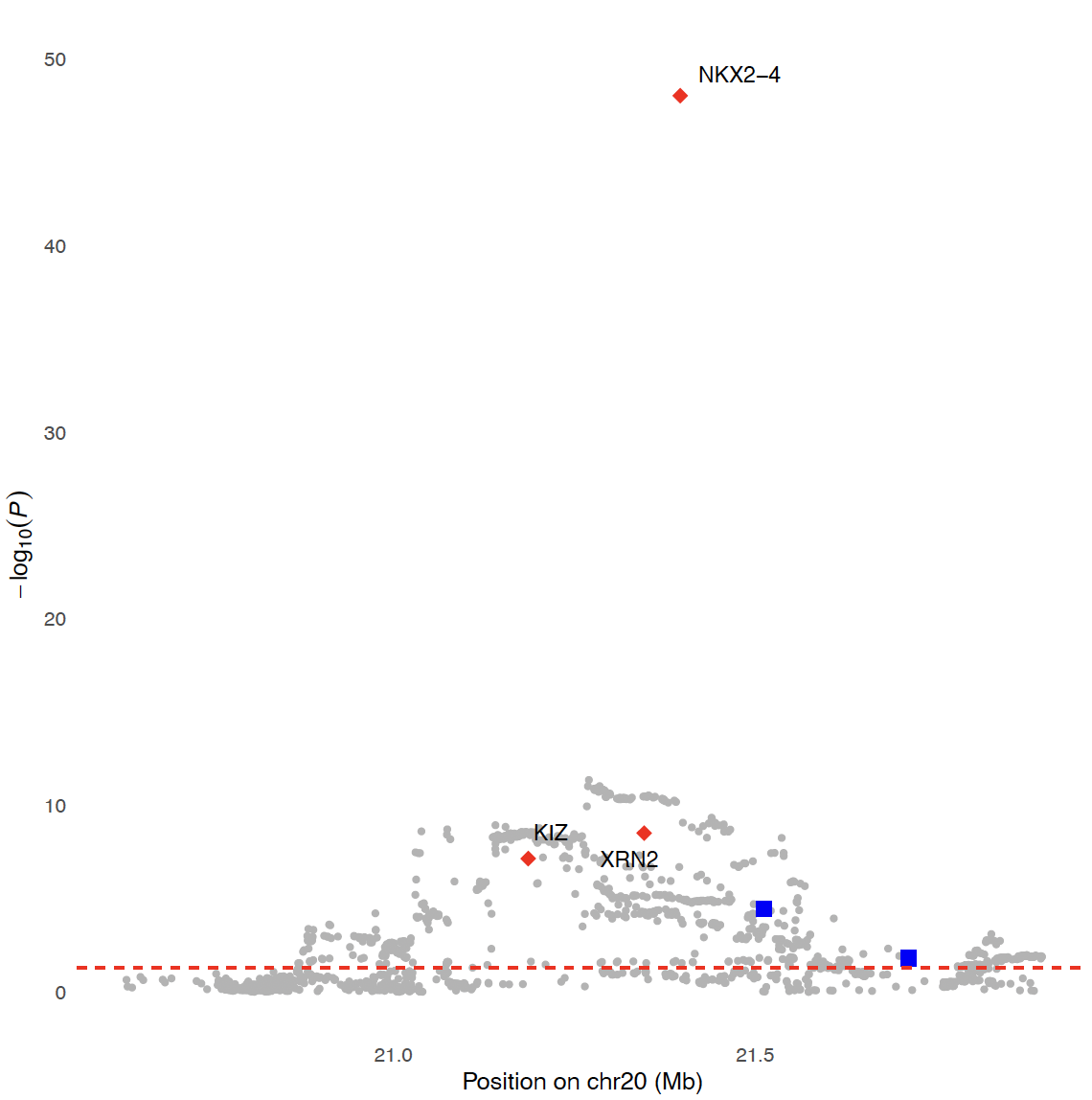

**Figure S11:** **GIFT results for basal ganglia at region chr1:42781877-43852772.** The gray circle represents the -log10(p-value) of each SNP from the GWAS, the blue square represents the -log10(OTTERS p-value) of each gene from the OTTERS TWAS, and the red diamond represents the -log10(p-value) of each gene from GIFT. The red dashed line represents the GIFT significance threshold of 0.05. All genes that were statistically significant in the TWAS are labeled.

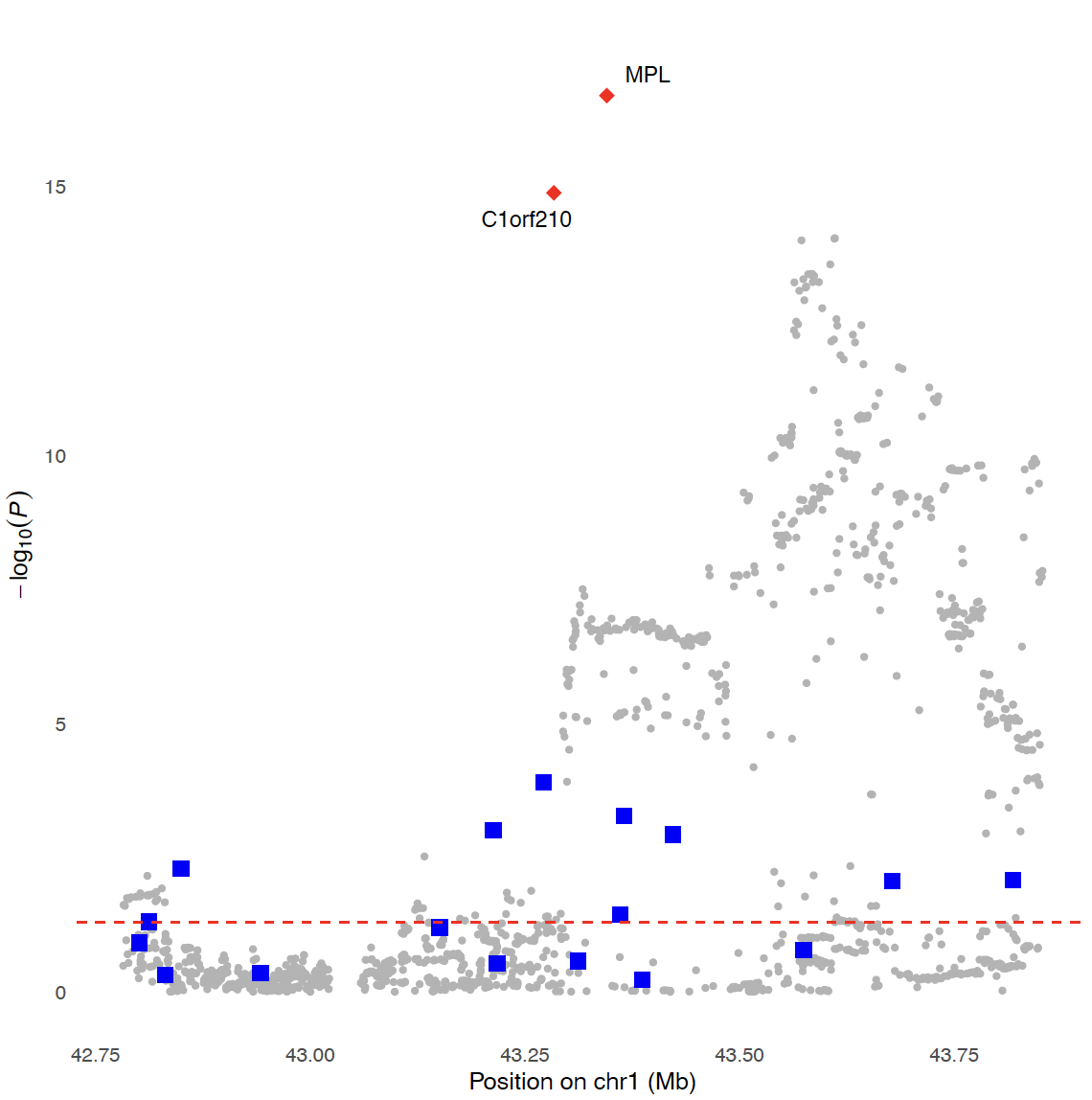

**Figure S12:** **GIFT results for cerebellum at region chr1:42645153-44496528.** The gray circle represents the -log10(p-value) of each SNP from the GWAS, the blue square represents the -log10(p-value) of each gene from the OTTERS TWAS, and the red diamond represents the -log10(p-value) of each gene from GIFT. The red dashed line represents the GIFT significance threshold of 0.05. All genes that were statistically significant in the TWAS are labeled.

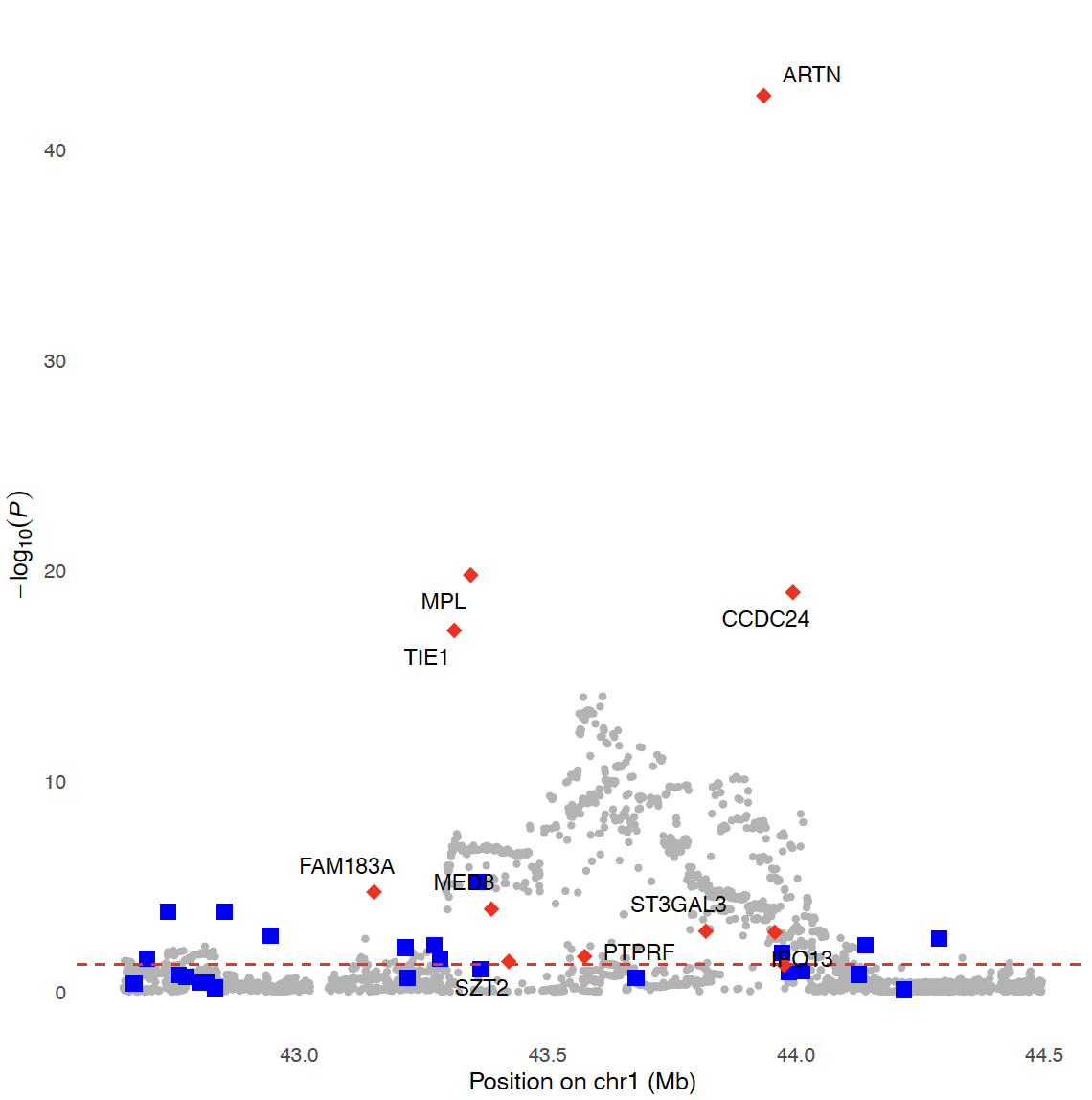

**Figure S13:** Protein-Protein Interaction (PPI) network analyzed by STRING. Nodes represent proteins. Colored nodes represent queried proteins and first shell of interactors. Edges represent protein-protein associations.

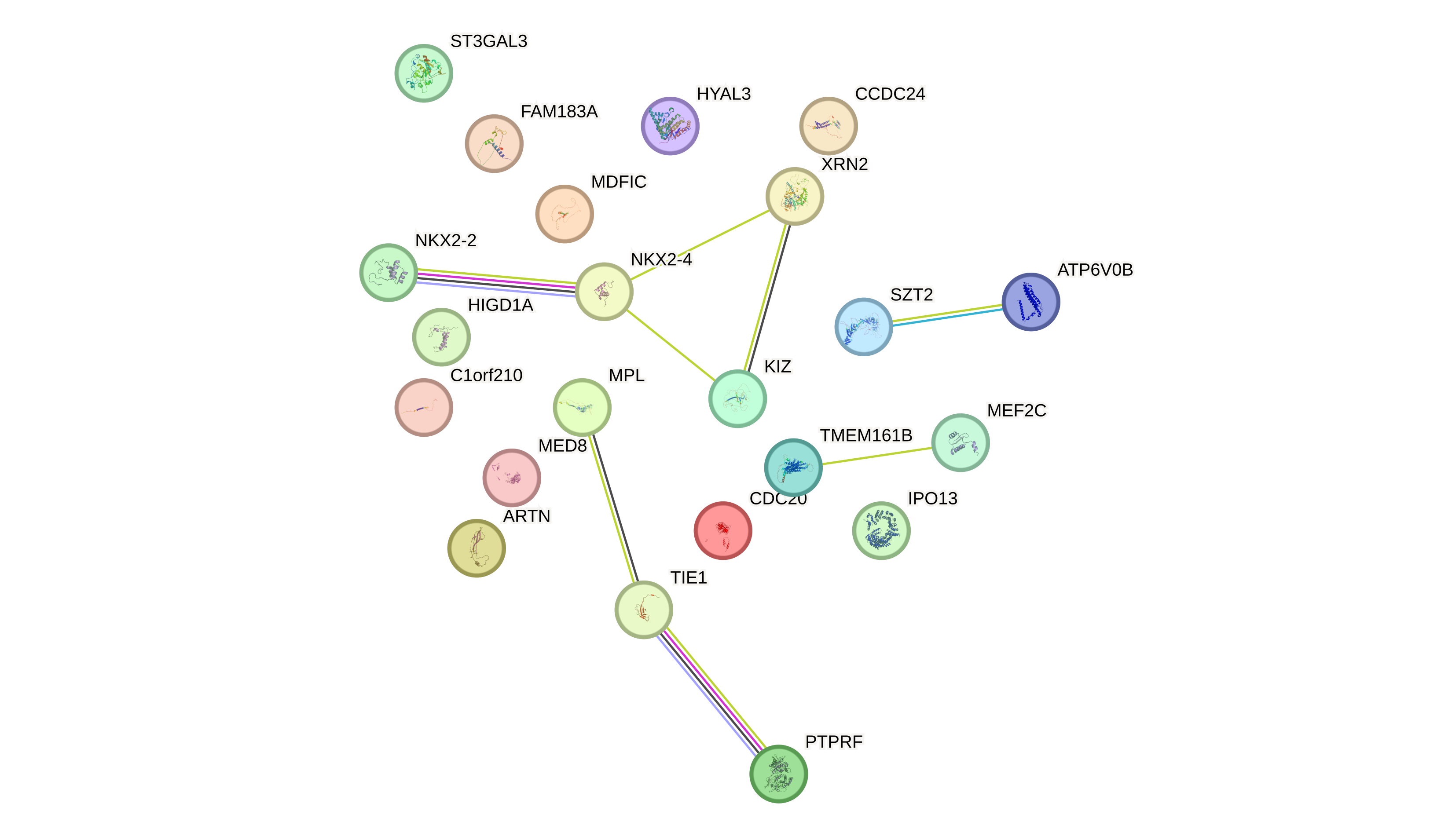

**Figure S14:** Enriched reference publications as analyzed by STRING. Significance threshold determined by FDR <0.05
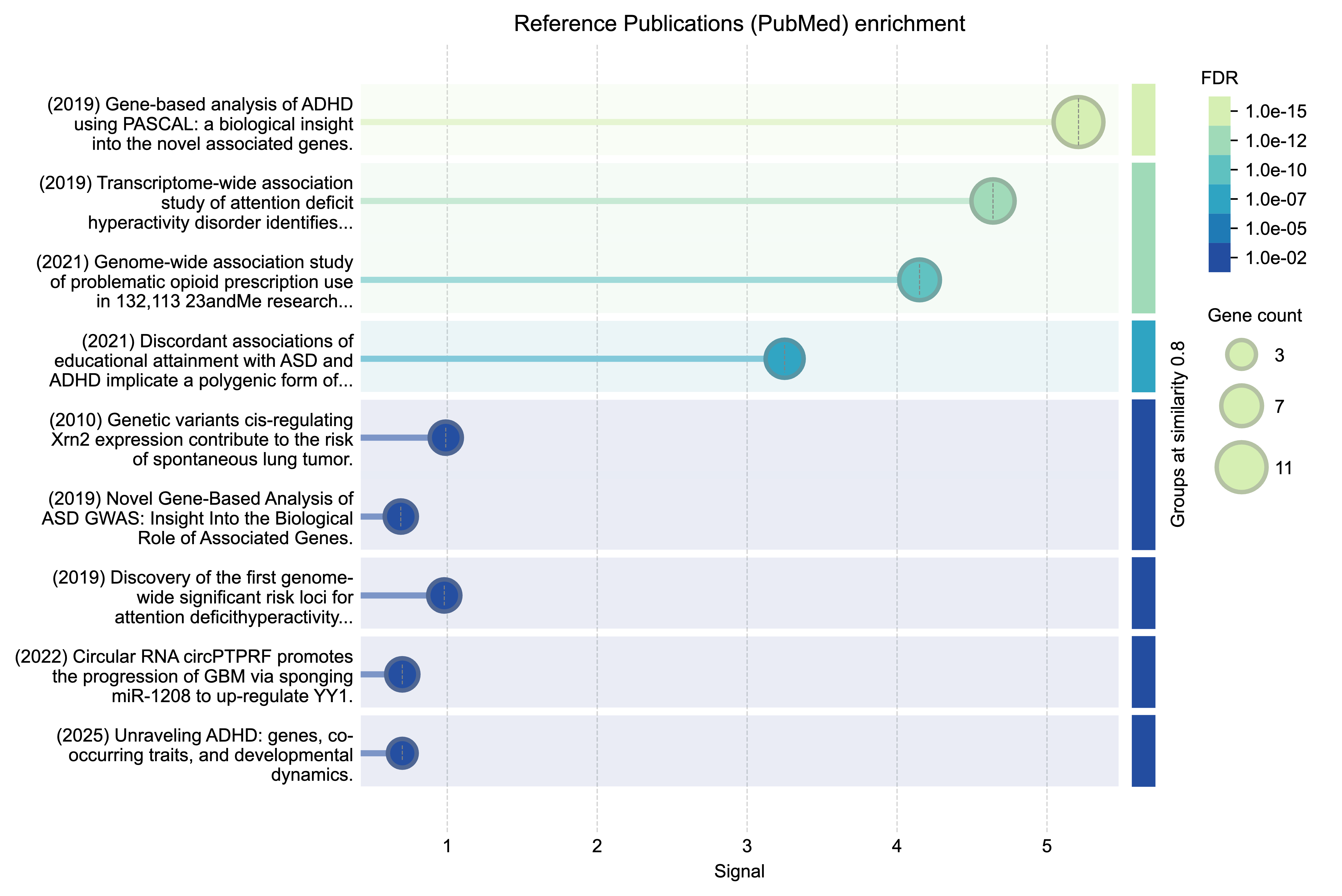
